# Comparative Analysis of the Application of Behavioural Insights of 33 Worldwide Governments on the Landing Pages of their COVID-19 Official Websites and their Impact on the Growth Scale of the Pandemic

**DOI:** 10.1101/2020.05.15.20103424

**Authors:** Paola Cecchi-Dimeglio, David Luu, Yann Cabon, Raphaëla Kitson-Pantano

**Author notes:** These authors contributed equally to this work (Paola Cecchi-Dimeglio, Raphaëla Kitson-Pantano). These authors also contributed equally to this work (David Luu, Yann Cabon).

## Abstract

The COVID-19 crisis has seen over a third of the world population locked down and this article has sought to understand human behaviour in response to a historical and unprecedented global pandemic. Through the analysis 18 behavioural mechanisms present on the landing pages of the websites of 33 institutional governments from March 1st til May 1st 2020 compared to the WHO data on the number of COVID-19 cases and deaths per million for each country, the authors show that a behavioural consensus was observed across all 33 countries and that Individual and Social nudges had no impact. Whilst the decisions in essentially every country on Earth, were taken with the same aim: to limit population movements and social life, two aggravating factors of the spread of the virus, only the environmental nudges effectively helped slow the virus growth scale. The authors explain the rationale behind these results and suggest that people seek information beyond governmental websites that they generally mistrust. They further suggest using Scientists as role models to encourage governmental website’s traffic and designing recursive nudges to increase the impact of individual and social interventions. Together with the new phases of the spread of the virus will come new rules and guidance. Public health policies need to address behavioural change of the population on a global scale in a more targeted manner and it is hoped that this paper will provide some insight on how to do so.

## II. Introduction

All human-beings on the planet share a 3 billion DNA code that differentiates them from other species; but the exact sequence of each, combined with individual life-experiences and behaviours makes every one of us unique. There are, on this planet, over 7 billion different humans! Understanding human behaviour has been the focus of many disciplines over the centuries…anthropology, sociology, psychology, evolutionary biology, human behavioural ecology and so on. And indeed, the human species not only displays an extraordinary range of ecological settings but also exhibits enormous behavioural diversity [1]. “People vary in their social, mating and parental behaviour and have diverse and elaborate beliefs, traditions, norms and institutions” [2]. There are some behavioural characteristics unique to some populations and absent in others and there are those that are common in all populations, but exhibit variation in their expression [3]. For example, whilst language is common to all humans, there are a plethora of different ones. Further there are behavioural differences observed within populations themselves, depending on age, gender, social status etc. such that individuals from a same population develop different roles, rights and responsibilities [4]. “Behaviour also varies within individuals over the course of their lifetimes [5]. Despite these behavioural differences, at time of writing, over a third of the world population was locked down. Over two billion people were successfully ordered and accepted to stay at home in order to mitigate an invisible and nevertheless deadly threat. This article has sought to understand human behaviour in response to a historical and unprecedented global crisis. Through the analysis of the behavioural interventions used by 33 governments on the landing pages of their official COVID-19 websites, the authors show that the observed human reactions to this unexpected challenge was very different to what had been anticipated and thereby shed light on the relevant nudges to implement as the crisis continues to unfold until a vaccine or a cure can be developed.

### The historical, unprecedented but expected COVID-19 pandemic

In 2006, Morse *et al* published an article entitled “*Next Flu Pandemic: What To Do Until The Vaccine Arrives?*” that stated that “experts believed the world was overdue for influenza pandemic” [6]. The COVID-19 pandemic is considered historical because very few of us today were alive when the last pandemic of similar scale (the Spanish flu) plagued the world. It is unprecedented because a pandemic in a globalized and interconnected world like the one we are currently living in has never occurred before [7]. However, this pandemic was expected and governments had prepared contingency plans. Nevertheless, Morse’s paper expressed the concern that “unless effective action against pandemic flu is taken now, we are in “dire straits,”” [8]. Indeed, as immunization was the cornerstone of countries’ strategy, with antiviral agents as a backup, producing and distributing a vaccine was estimated to take at the very least 4 to 6 months. The contingency plans therefore focused on nonpharmacological interventions, such as hand washing, “respiratory etiquette,” face masks, school closures, and social distancing or isolation. However, the effectiveness of these measures was based on those used in 1918 and remained poorly understood [9].

As the COVID-19 virus emerged in December 2019, sparking an epidemic of acute respiratory syndrome in humans in China, within three months, the virus had spread to more than 118,000 cases and caused 4,291 deaths in 114 countries, leading the World Health Organization to declare a global pandemic [10]. (*The number of cases, deaths and the dates detailed in this paper are the official numbers accessible by the authors at the time of writing. As the crisis continues to unfold and the COVID-19 disease continues to spread, these numbers and dates may be subject to change*.) On April 20^th^, 2020, the total number of known cases was greater than 2.36 million worldwide [11]. By then almost a third of the world’s population was under some form of lockdown. There is a lot of subject matter related to this COVID-19 crisis that is, and will remain, hard to assess (speed of transmission, likelihood of survival, economic consequences etc.). An important issue is how to communicate uncertainty and share information when so little is actually known. The behavioural response to COVID-19 is unavoidably collective. Each person’s chance of contracting the virus depends not only on their own behaviour, but also on the behaviour of their fellow citizens [12].

This paper proposes to review the published literature on the successful leveraging of behavioural insights during past epidemics and on the data available at time of writing on the current pandemic. Whilst countries response worldwide was predominantly national, how global leaders embraced the diversity of the behavioural insights specific to their own citizens to design the landing page of their governmental institutions’ website will be assessed. It will be shown that far from expectations, the response to the sanitary crisis was very consensual with countries implementing 80% of the behavioural insights observed. The research shows that beyond this consensus, the individual and social nudges had no impact on the number of cases and deaths of COVID-19 observed and instead it was the environmental interventions that had a significant impact. What are the drivers that explain these results will be later discussed.

### The relevance of Behavioural Science insights in fighting a global pandemic

Understanding a global population’s response to a worldwide pandemic such as COVID-19 is not straightforward and requires rigorous analysis. Below are four putative applications of behavioural science that are relevant to understanding this. ^1.^Behavioural science can not only be used as a powerful tool to understand humans, ^2.^it is key to drafting impactful and targeted messages to support crisis communication techniques and to ensure that what is conveyed is heard and perceived as intended. ^3.^Behavioural science insights can help leverage the feeling of belonging and ^4.^finally can be used to direct human behaviours toward outcomes [13].

^1.^Levering insights from behavioural science requires not only to focus the attention on the decision making and behaviour of citizens (the end users) but also to target the behaviour of those at the source of the process, the policy makers, who communicate down to the citizens, thereby addressing those people who design the process [14]. Indeed, whilst “end-use behaviour can determine what happens in a situation, design behaviour often determines the situation itself” [15]. Behavioural science uses multiple behavioural levers such as social influence, convenience, prompts, and cues [16] all tools that can impact what is known as the “Behaviour Change Wheel” that suggests that change is only possible if capability, opportunity and motivation are aligned. “Individuals have to be psychologically or physically able to undertake the behaviour, the environment that surrounds them needs to facilitate the behaviour, and their own mental processes need to energise and direct the behaviour” [17]. Further, research demonstrates that behaviour change is more likely when the behaviour is made Easy, Attractive, Social and Timely [18].

^2.^Behavioural Science gives relevant insights on how best to address populations with life-changing news. Clarity in the messages that the governments convey is essential to ensure readability and understanding. Research has shown that clarity and certainty about timelines are both important [19] and that “messages linked to disgust tend to be more effective than messages that communicate social norms” [20]. Further, plans are easier to follow if they are time-specific and intentional, rather than general aspirations [21]. Co-operation is increased by clear statements, articulated by leaders and repeated by others, of a desired collective behaviour that is in the group interest. This can enhance trust, establish social norms and encourage individuals to commit to the behaviour [22]. Consequently, behavioural science helps design messages that elicit emotions other than fear such as ‘empathy appeals’ that can have positive impacts on behaviour change [23]. Making communication sensitive to the demographics of the intended recipient helps people to feel that society is more prepared. Further, behavioural science has shown that catchy phrases or mnemonics help children and families retain important public health information [24]. The content of the message needs to be further supplemented by the form the message takes. Bright infographic designs are more efficient at communicating a step-by-step procedure for thorough hand-washing and the best place to display this hand-washing procedure and instructions increases compliance to high quality hand-washing. Communication clarity increases the efficacy of taking action. When a crisis as devastating as the COVID-19 crisis hits, there is necessarily an information asymmetry between what the governments are receiving as data from scientists, experts and other governments worldwide and the information that can be communicated down to the population. In addition to the way the message should be conveyed, the way it will be perceived needs to be accounted for when drafting the message.

^3.^Humans are social creatures and their desire to connect to people goes beyond wanting to interact with family, colleagues and friends. A fundamental human need is the feeling of belonging. Studies have shown that a sense of belonging is associated with numerous beneficial outcomes, whereas a broken sense of belonging increases the risk for psychological and physical dysfunction. Behavioural science has shown that community-spirited behaviour can be enhanced through communication, group identity and punishment [25]. Leaders who are perceived as ‘one of us’ and as ‘working for us’, rather than for themselves or for another group, tend to gain greater influence [26]. Building a strong sense of shared social identity can help coordinate efforts to manage threats and foster in-group commitment and adherence to norms [27]. The more people feel part of a group or community response, the more likely they are to make a selfless contribution. This finding is particularly true of responses to threats, which generate a stronger public response when framed in group rather than individual terms [28]. Co-operation is more likely when there is transparency about individuals’ contributions and punishment for those who do not pull their weight [29].

^4.^Finally, insights from behavioural science can help change people’s behaviours. Habits are a positive drive in our lives, designed to free up our minds to concentrate on other matters, “by definition, they operate mostly outside conscious awareness and are hence hard to break” [30]. Even in acute healthcare environments, attempts to improve hand hygiene and other infection control behaviours through education and awareness have limited and short-term impacts [31]. Behavioural science insights help develop interventions that attract attention and make compliance convenient [32]. People’s behaviour is influenced by social norms such as what they perceive that others are doing or what they think that others approve or disapprove of [33]. Further, individuals struggle to perceive risks accurately and distort probability when making decisions, with substantial differences [34]. Appealing to fear leads people to change their behaviour if they feel capable of dealing with the threat, but leads to defensive reactions when they feel helpless to act [35]. Instead, exposure to celebrity advocacy messages can impact issue engagement, as long as the issue has previously been perceived as important and the celebrity has a favourable sympathy capital [36]. Research has shown that exposure to celebrities through the media can have an important influence on the public’s health-related attitudes, beliefs, and behaviour [37]. Involvement with a celebrity through media exposure is an important mediating variable in persuasive communication, and celebrities can effectively endorse health-related messages. When health rises up the priority ladder at the expense of all other considerations, as is the case in a Public Health crisis such as the COVID-19 pandemic, this leads to the increase of difficult choices to be made. The average person makes an eye-popping 35,000 choices per day and in the time of crisis the ones that citizens will need to make will have an impact on their life but also on society as a whole and on the global economy. Insights given by behavioural science will be impacted by the population the study has been conducted on and the cultures of those populations. Research has shown for example that Western European and North American cultures that endorse individualism and tend to positively value the expressivity of the self through kissing, hugging, and direct argumentation. Other cultures share a stronger commitment to collectives such as country, tribe and family the priority given to obligations and duties in Asian societies may motivate individuals to remain committed to social norms while suppressing personal desires [38]. Behavioural science can help ensure that reactions are based on logic rather than reactance. Behavioural Science has shown that voluntary quarantine is instead associated with less distress and fewer long-term complications. Further it is important to emphasize the altruistic choice of self-isolating.

### What behavioural science tells us about the response?

Faced with an outbreak such as COVID-19, it is important to understand what influences people’s behaviour in order to decrease the likelihood of infection, transmission, and disease severity. This understanding can inform communication strategies aimed at minimizing the impact and spread of the disease [39]. “Protective behaviours carried out in response to an influenza pandemic can be broadly classified into three types: preventive, avoidant, and management of disease behaviours” [40]. Hand washing and the use of alcohol hand sanitizers, coughing or sneezing into the elbow or the sleeve, cleaning surfaces are examples of preventive behaviours. Stay at home orders, social distancing and imposed teleworking are classed in the avoidant behaviours’ category. Management of disease behaviours include widespread testing and taking antiviral medication. These behaviours may be the remit of the individual or decreed by law [41]. Since a vaccine and or a treatment were not rapid enough solutions to the COVID-19 pandemic, governments worldwide focused on slowing down the viral transmission in order to limit the number of severely sick in hospitals and be able to attend to their medical needs in the most efficient way possible. In order to achieve this, significant shifts in behaviour were required but various aspects of social and cultural contexts influence the extent and speed of behaviour change [42]. A recent study based on a social media survey translated into 69 languages and addressing over 100 000 volunteers in 175 countries reported that 91% of respondents did not attend any social gatherings, 89% washed their hands more frequently and 93% declared that they would have immediately informed people around them if they had experienced COVID-19 symptoms [43]. Despite these very encouraging results, the virus continues to spread at an impressive pace, decimating the world population in its tracks.

In order to understand what drives behavioural changes and how efficient these changes actually are, current research has focused on seven: (1) Hand Washing; (2) Face Touching; (3) Entering and Coping with Isolation; (4) Encouraging Collective Action; (5) Avoiding Undesirable Behaviour; (6) Crisis Communication; and (7) Risk Perception [44]. The importance of making handwash solutions readily available and placing them in full view together with impactful signage in central locations has been shown to be an effective contributor to encouraging a more rigorous and frequent hand washing habit than usual [45]. Interestingly, to the best of the authors knowledge, there has been no evidence of any behavioural insight having been successfully leveraged to reduce face touching. Similarly, companies that already had remote work guidelines in place were more prepared to support their employees and address the issues linked to isolation such as distress and mental health problems. Because the conditions of telework under the COVID-19 crisis were somewhat different to a “normal” remote day at home…due to the length of the stay at home order and the perceived imprisonment of the quarantine, companies that actively connected with their employees on a regular basis were more successful at ensuring continued efficacy and productivity of their staff. Planning for the effects of social isolation can help individuals to cope. Plans may be put in place to remotely engage with social networks, via phone and video calls, or social media [46]. Research showed that Public-spirited behaviour was most likely when there was clear and frequent communication, strong group identity, and social disapproval for those who don’t comply [47]. This was evidenced by the social media survey mentioned above that showed that 90% of respondents believed social distancing measures were effective and 70% felt that risky behaviours should be financially punished [48]. Research shows that authorities often overestimate the risk of panic [49], and whilst in some populations, for no logical reason, people overpurchased specific goods such as toilet paper, this was not the most common reaction. Further, this compulsive purchasing has been shown to be a psychological response to the need of being in control of something. “When deciding whether to engage with proposed health solutions, people consider not only their susceptibility to the threat and its severity, but how effective they perceive the solution to be and the nature of the required behaviour” [50]. “One of the most effective arguments to promote compliance with isolation is that self-isolation in response to symptoms is the best way for all of us to prevent infecting each other” [51]. The impact of communication on the behaviour of people together with the perception of risk will be further addressed. Indeed, these are only two of the factors that can impact behavioural changes together with age, gender, ethnicity, education level, social norms to name but a few.

Research shows that whilst in some populations there are behavioural changes associated with age, the pattern of findings for age is not straightforward. The balance of evidence shows that increasing age is associated with a greater chance of carrying out behaviours, but overall, the results are inconclusive [52]. Gender however has a strong impact on behaviour and research shows that women are consistently more likely than men to carry out the behavioural changes imposed in a time of pandemic [53]. “There is insufficient evidence to draw any firm conclusions about associations between ethnicity and pandemic-related behaviours” [54]. More educated people are likely to take protective and avoidant behaviour and a relationship has been found between avoidant behaviours and a higher perceived severity of the pandemic [55]. Social pressure has been shown to be associated with mask wearing and belief that the authorities are open with their communication is likely to impact the implementation of precautionary behaviours [56].

This article proposed to address the research question “*To what extent did the use of behavioural insights by governments on the landing page of their official COVID-19 websites affect the epidemic growth scale of COVID-19?*”. To do so the landing pages of 33 official COVID-19 websites were analysed as described below.

## III. Materials and Methods

### Data sets and parameters used

The starting point for the dataset used herein is the landing pages of the websites of 33 institutional governments. The authors declared no potential conflicts of interest with respect to the research, authorship, and/or publication of this article. Further the WHO data was used in order to access the number of COVID-19 cases and deaths per million for each country and on specific dates [57]. (*The dates the data was accessed were 1^st^ March, 15^th^ March, 1^st^ April, 15^th^ April and 1^st^ May 2020 for 33 countries: Austria, Belgium, Bulgaria, Croatia, Cyprus, The Czech Republic, Denmark, Estonia, Finland, France, Germany, Greece, Hungary, Iceland, Ireland, Italy, Latvia, Lichtenstein, Lithuania, Luxembourg, Malta, The Netherlands, Norway, Poland, Portugal, Spain, Sweden, the UK, Romania, Slovakia, Slovenia, Switzerland and the USA*). The complete list of the websites accessed and the date when these were accessed can be found in the supporting documents.

For every country, the “total cases of COVID-19” curve was defined by an exponential spread of the disease at the start of the epidemic, followed by the stabilization changes seen in individuals or groups and then an exponential decrease of the curve as the spread of the disease progressively slowed down. In order to estimate the epidemic evolution as well as the growth, a logarithmic equation fitting the “epidemic cumulative cases” curve of the COVID-19 cases and deaths adjusted per million inhabitants was used. Adjustment were carried out in order to remove the effect of the country-size demographic relation with the epidemic spread.

In order to address the research question “*To what extent did the use of behavioural insights by governments on the landing page of their official COVID-19 websites affect the epidemic growth scale of COVID19?*”, the data was adjusted to account for the six dimensions of national culture of Hofstede *et al* [58]. The application of this research is used worldwide in both academic and professional management settings [59]. Their model suggests that cultural dimensions represent independent preferences for one state of affairs over another. Because we are all human and simultaneously all unique, cultural dimensions distinguish countries (rather than individuals) from each other. Further, the country scores on the dimensions are relative and therefore culture can only be used meaningfully by comparison. The model consists of the following six dimensions:

#### 1. Power Distance Index (PDI)

This dimension addresses the extent to which the inequal distribution of power is accepted and expected by a population. A high level of PDI demonstrates that people accept the established hierarchical order and therefore do not question the fact that everyone has a set place that is determined. Inversely a low level of PDI means the population seeks to distribute power and demands an explanation for the lack of equality [60].

#### 2. Individualism Versus Collectivism (IDV)

High levels of IDV indicate a society driven by the feeling of Individualism. This is often associated with a loosely-knit social framework in which people care little for anyone beyond themselves and their immediate families. A low level of IDV is indicative of Collectivism, and a tightly-knit framework in which taking care of relatives is considered the norm and family loyalty is important. The difference between individualism and collectivism can be simplified to the dichotomy between “I” and “we” [61].

#### 3. Masculinity Versus Femininity (MAS)

Populations that exhibit a preference for achievement, heroism, assertiveness, and material rewards for success demonstrate a high level of MAS and are generally more competitive. A low level of MAS instead indicates femininity defined by cooperation, modesty, caring for the weak and quality of life. This results in decisions being made in a more consensual fashion [62].

#### 4. Uncertainty Avoidance Index (UAI)

A high level of UAI reflects the unease of a society with the concepts of uncertainty and ambiguity. To what extent does a population accept that the future cannot be controlled is revealed by the level of UAI. High levels of UAI are rooted in codes of belief and behaviour, and representative of an intolerance towards unorthodox behaviours and ideas. A low level of UAI indicates that the population cares more about practice rather than principles [63].

#### 5. Long Term Orientation Versus Short Term Normative Orientation (LTO)

Not all societies welcome change and perceive it as a sign of progress. Populations that exhibit a low level of LTO tend to be rooted in their past and emphasize their traditions and norms as they anticipate change with suspicion. Inversely, a high level of LTO is representative of a pragmatic approach. Populations with high LTO embrace the future and its challenges, prepare for it and educate the young in that respect. In business terms, this is known as the short term versus long term dichotomy [64].

#### 6. Indulgence Versus Restraint (IVR)

Populations that score high for IVR tend to be more relaxed and encourage free gratification, the enjoyment of life and fun. Inversely, populations that score low on IVR are more focused on strict regulations and the emphasis of social norms [65].

### Method carried out

Three levels of analysis were carried out: the individual level, the social level and the environmental level. For the individual level, two domains were analysed: 1 behaviour and 2. well-being and mental health. For the social level, the community spirit behaviour domain was assessed and for the environmental level, two further domains were 1. crisis communication and 2. risk perception. For each of the domains, actions were dissected: two for the behavioural domain at the individual level (1. handwashing and 2. face touching), and one for the well-being domain at the individual level, (isolation). For the social domain the action was incentives and for the environmental level, for both domains, the actions were the cues. In total, 18 mechanisms were observed, assessed and compared across the 33 countries. When the mechanisms were present on the website the value “1” was assigned, when they were absent, the value “0” was assigned.

The websites were accessed at a date t = 0 (every two weeks from March 1^st^ til May 1^st^) but the evolution of data 15 days beforehand and 15 days before that, all the way back to the 1^st^ of March was assessed in order to derive cumulative data sets and to identify the order in which the information came in.

Further, the authors wish to emphasise a major caveat of this study which is that whilst the results hold true at the time of writing, because the COVID-19 is an ever-evolving crisis that is ongoing, the results may differ as we progress in the crisis. Further, there are some websites that do not mention the date of their updates and don’t update their information regularly. This has been accounted for in the mathematical models described below.

An additional caveat that makes epidemic data complex to analyse is that they are non-linear, time correlated and the date of first infection and following spread depend on a plethora of external factors. The initial exponential growth scale of an epidemic is an important measure of the virulence of the epidemic and is also closely related to the basic reproduction number.

Logarithmic equations are commonly used for these types of analysis and consequently were used to model the growth scale of the disease [66]. Governments’ use of behavioural insights on the landing pages of their official COVID-19 websites were analysed to assess whether there was an effect on the epidemic spread of the disease. The model was adjusted by using the Hofstede scale in order to account for the impact of the different country cultures.

The mathematical model used looked at the epidemic growth. Epidemic curves are time series data of the number of cases per unit time. Common choices for the time unit include a day, a week, a month, etc. It is an important indication for the severity of an epidemic as a function of time. We used the common approach of a logistic model to assess the population growth dynamics. The function of the model is as follows:

Let us define the cumulative number of cases *Y* over time *t*, with a growth scale *s* and a maximum case possible *A*.

The population evolution can be modelled with the following equation as

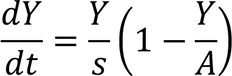

This definition is compliant with the limit hypothesis:

­ 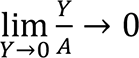
­ 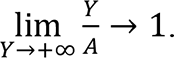

The parametric form of logistic equation is therefore

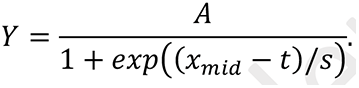

The parameter *A* is the asymptotic value of the logistic equation.

*x_mid_* is the inflexion point of the epidemic evolution.

*s* is the scale parameter, where the higher the value, the lower the epidemic growth.

Estimation of this nonlinear system was done with R 3.6.1 software and the SSlogits function. The parameter’s initial value was automatically computed, and the parameters were estimated with a Gauss-Newton algorithm [67].

The aim of this study was to assess epidemic growth per country. The parameter conserved for the statistical analysis was therefore *s*

To assess the efficacy of behavioural insights and country specifications over the epidemic, growth scale was computed with a linear quadratic regression. Adjustment on Hofstede’s scale was done to adjust behavioural insights regarding population behaviour using a linear model on the criteria (*s*). These models included Hofstede variables as interactions of the linear model. Interactions represent the combined effect of two variables over the studied one.

Data was presented as mean (μ_x_) and standard deviation (σ) of the estimate. Estimates were computed with the maximum likelihood framework. Significance of the model was taken at *p* < 0.05. Normality was assessed with residual plots due to the size of the sample.

## IV. Results

In order to compare accurately the use by governments of behavioural insights on the landing pages of their official COVID-19 websites, the data was adjusted to account for cultural differences [68]. The six dimensions of national culture of Hofstede *et al* were used. Table 1 shows for each of the dimensions the mean result for all countries where data was available (N-NA’s) together with the minimum and maximum values for each dimension. Table 1 also shows the mean use of individual, social and environmental factors as well as the mean growth scale.

**Table 1:**
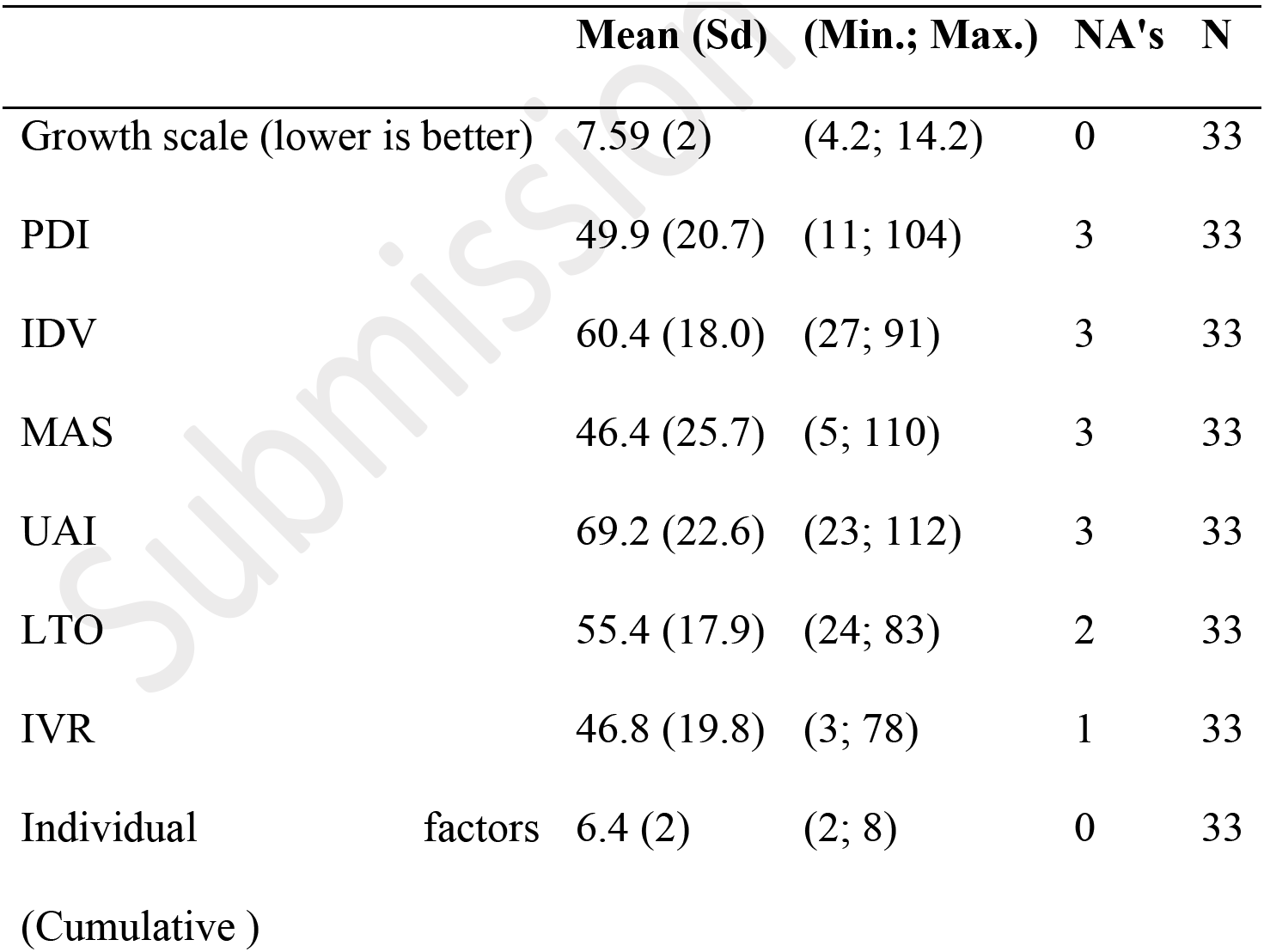
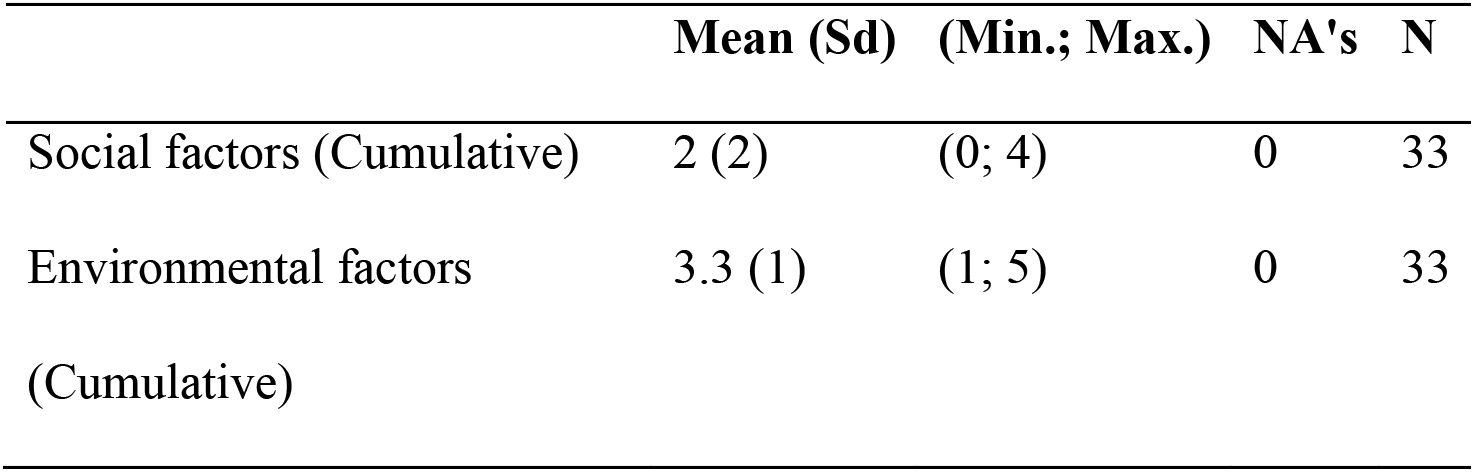
Descriptive Quantitative Data (I)

The results are based on the growth scale estimate with the logarithmic equation for each country as presented previously. A high value of growth scale means a low spread of the disease in the population and a low value of growth scale means a higher spread of the disease.

For each of the six dimensions of national culture, looking at the mean values gives an appreciation of the cultural biases of the countries under analysis. For instance, a PDI mean value of 49.9 indicates that half of the countries accept and expect that power is distributed unequally. The top six countries accepting a hierarchical order, in which everybody has a place, are unsurprisingly Slovakia, Romania, Croatia, Slovenia, Bulgaria and the Czech Republic. These eastern European countries were until recently a part of the USSR and whilst their opening and joining of the EU has broadened their horizons, the cultural biases are still very present.

The mean value for IDV is 60.4 indicating that most countries under analysis in this study are majoritarily more individualistic, with a preference for a loosely-knit social framework in which individuals are expected to take care of only themselves and their immediate families, than collective. The USA and the UK are the top two countries exhibiting this cultural trait.

Whilst almost half of the countries display more masculinity and therefore are more competitive, the five countries where a more consensus-oriented society (femininity) is the highest are Denmark, the Netherlands, Latvia, Norway and Sweden.

As could be expected in view of the context, the highest mean value is for the Uncertainty Avoidance Index dimension which reasserts that the majority of countries were uncomfortable with the uncertainty of the COVID-19 pandemic.

Further table 1 shows that of the 9 Individual factors that were analysed, the majority of countries implemented on average 80% of them (mean = 6.4 out of maximum 8). Note here that the maximum value is 8, not 9, because for one of the factors, all 33 countries implemented it and it was therefore removed from the comparison between countries. In contrast, only half of the social factors mechanisms on average were implemented. Lastly, on average 66% of the environmental factors were implemented across the 33 countries.

Table 2 is a more detailed quantitative analysis of the usage by governments of behavioural insights on the landing pages of their official COVID-19 websites of each of the 18 mechanisms observed.

**Table 2:** Descriptive Quantitative Data (II)

| Variables Factors | Yes<br>(%) | No<br>(%) | N |
| --- | --- | --- | --- |
| **Individual Factors (Behavioural change)** |  |  |  |
| - Handwashing & Education | 82% | 18% | 33 |
| - Handwashing & Information | 88% | 12% | 33 |
| - Handwashing & Feeling eliciting “disgust” | 0% | 100% | 33 |
| - Handwashing & Feeling eliciting “shift in social norms” | 100% | 0% | 33 |
| - Encourage a specific change in social acceptability* | 82% | 18% | 33 |
| - Encourage self-isolation | 100% | 0% | 33 |
| - Provide help with an COVID19 support Hot line | 88% | 12% | 33 |
| - Provide help in planning with isolation coping | 85% | 15% | 33 |
| - Provide guidance for maintain aspects of routine | 18% | 82% | 33 |
| *\*e.g. perhaps that scratching with a sleeve is fine, just as it is now advised to sneeze or cough into the elbow or upper arm rather than the hand* |  |  |  |
| **Social Factors (Incentives change)** |  |  |  |
| - Foster cooperation by engaging population via communication medium (SMS/ push notifications) | 27% | 73% | 33 |
| - Transparency & individuals' contributions | 64% | 36% | 33 |
| - Punishment "financial fine" | 48% | 52% | 33 |
| - Inclusive language ("We" versus "I" or "you") | 61% | 39% | 33 |
| **Environmental Factors (Social Information Processing Cues change)** |  |  |  |
| - Cues eliciting emotions other than fear such 'empathy appeals', 'responsibility appeals' | 100% | 0% | 33 |
| - Cues eliciting protection of vulnerable group (older, pregnant women...) | 42% | 58% | 33 |
| - "Availability heuristic" | 64% | 36% | 33 |
| - "Hindsight bias" | 70% | 30% | 33 |
| - "Framing effect " | 58% | 42% | 33 |

Two results are particularly significant. First, not one country used the mechanism of disgust to encourage handwashing. All 33 countries encouraged handwashing through alternative mechanisms but none resorted to making their citizens feel disgusted by the fact of having dirty hands contaminated with the virus. A second result that is common to all countries is that all 33 countries used social norms to encourage handwashing. Similarly, all 33 countries encouraged self-isolation and designed their websites to elicit emotions such as empathy and responsibility. None of the websites assessed among the 33 countries used mechanisms to elicit the feeling of fear or shame [69].

Whilst 29 out of 33 countries all provided information about the importance of handwashing and provided a hotline help number, the Netherlands did not provide any information about the significance of handwashing and Slovenia did not provide a hotline access. Further, Lithuania, Cyprus and Croatia provided neither.

Interestingly, the six countries that did not provide any education on how to wash hands appropriately to minimize contamination with the COVID-19 virus, were the same six countries whose Public health authorities did not encourage a specific change in social acceptability, perhaps that scratching with a sleeve is fine, just as it is now advised to sneeze or cough into the elbow or upper arm rather than the hand. These countries were Austria, Croatia, Cyprus, Finland, Greece and the Netherlands.

All in all, three countries stand out through the fact that they neither promoted education, information or any feeling of disgust in handwashing and solely relied on the social norms’ aspect of the mechanism. These were the Netherlands, Cyprus and Croatia.

Additionally, Austria, Croatia, the Czech Republic, Denmark and Lithuania were the five countries that did not provide help to support the coping of self-isolation through social networks, phone and video calls or social media.

In particular Lithuania and Croatia stand out because they were the two only countries that did not provide help to support the coping of self-isolation through social networks, phone and video calls or social media or provide a hotline access.

Only six countries (Iceland, Ireland, Luxembourg, Norway, Switzerland and the USA) out of 33 provided guidance for maintaining aspects of routine during self-isolation and two thirds of those countries, all but Iceland and the USA, also promoted communication enhancing cooperation (via what’s app number) together with five other countries (UK, Sweden, Portugal, Liechtenstein and Germany). But for both these mechanisms the countries who used these on their websites were by far a minority.

Interestingly, Norway and Switzerland adopted the exact same behaviours for all 18 mechanisms. The USA did the same as well with the exception of one mechanism…the USA did not use the communication enhancing cooperation (via what’s app number). All the others were the exact same as for Norway and Switzerland.

At the individual level, six countries used the exact same mechanisms for every action under every domain. These were Bulgaria, Estonia, Hungary, Latvia, Romania and Slovakia. Further Estonia and Hungary were also aligned on the exact same mechanisms for every action under every domain of the environmental level.

Another five countries used the exact same mechanisms for every action under every domain at the individual level, although different from the previous six. These were France, Italy, Malta, Portugal and Spain.

Next, how the cumulative use by governments of behavioural insights such as individual factors, social factors and environmental factors on the landing pages of their official COVID-19 websites affected the growth scale of the pandemic was assessed. Table 3 shows these results.

**Table 3:** Epidemic scale growth with factors analysis.

| Factors & Epidemic scale growth |  | Estimate | p-value |
| --- | --- | --- | --- |
|  |  | Mean (Sd) |  |
| **Environmental factors** |  |  |  |
|  | (Intercept) | 9.34(0.72) | 0 ** |
|  | Environmental | -0.54(0.2) | 0.01* |
| **Individual factors** |  |  |  |
|  | (Intercept) | 6.92(1.36) | 0 ** |
|  | Individual | 0.1(0.21) | 0.64 |
| **Social factors** |  |  |  |
|  | (Intercept) | 7.99(0.51) | 0 ** |
|  | Social | -0.22(0.21) | 0.29 |

Table 3 shows that the use of individual factors and social factors had no effect on the epidemic scale growth (p = 0.64 for individual factors and p = 0.29 for social factors). However, the use of environmental factors affected the growth scale (epidemic logarithmic equation growth). This suggests that countries applying more environmental behavioural insights demonstrate a lower and slower growth of the COVID-19 pandemic.

This is further illustrated in Figure 1 that shows how the growth scale of the COVID-19 pandemic is impacted by the use of environmental factors behavioural insights by governments on the landing page of their official COVID-19 websites.

**Figure 1:**
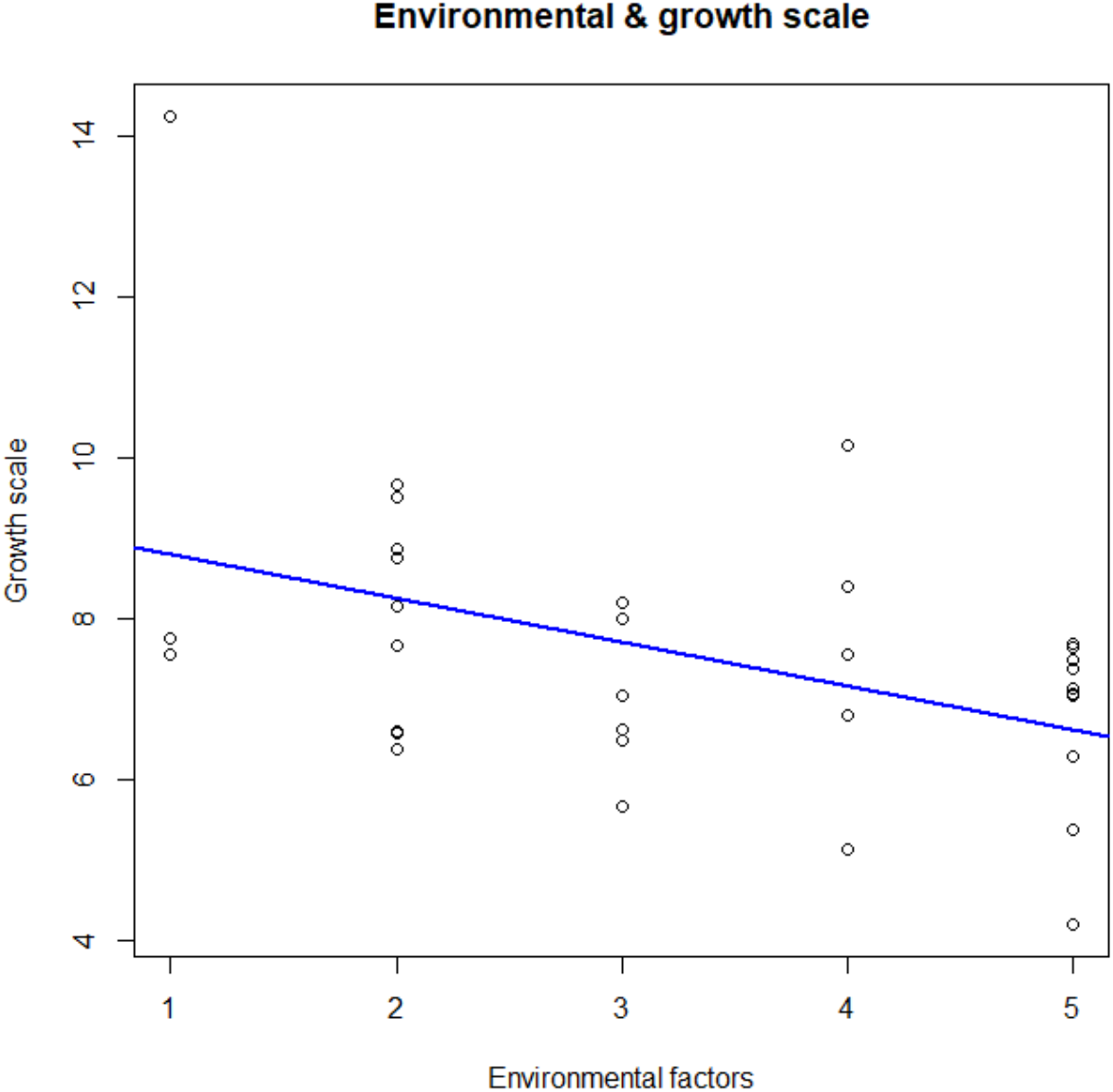
Environmental & Growth Scale

The horizontal axis represents the cumulative number of environmental factors behavioural insights used by governments on the landing page of their official COVID-19 websites. The growth scale is derived from the logarithmic equation model. The higher the value of the growth scale the slower the disease was spreading and the lower the number of cases. Inversely a low value of growth scale means a high increase of the spread of the disease and of the number of cases. The blue line represents the linear estimate of the relation between growth scale and environmental behaviour application. What is observed is that the growth scale decreases per 0.06 per environmental behaviour applied. Environmental factors behavioural insights application tend to reduce the spread of COVID-19 and the increase in number of cases.

Whilst every one of the 18 mechanisms was adjusted to account for the six dimensions of Hofstede, only one mechanism showed a significant result (p<0.05) as shown in Table 4.

**Table 4:** Growth scale adjusted on *IVR*.

|  | Estimate |  |
| --- | --- | --- |
|  | Mean (Sd) | p-value |
| (Intercept) | 11.42(1.31) | 0 ** |
| “hindsight bias” – No | -4.13(1.54) | 0.01* |
| IVR | -0.06(0.03) | 0.02* |
| “hindsight bias” – No & IVR | 0.06(0.03) | 0.05. |

Table 4 shows that when the mechanism “hindsight bias” is not used by governments on the landing page of their official COVID-19 websites a higher epidemic growth can be observed in the country. –4.13 is representative of a higher epidemic growth in the country.

When the Hofstede variable “IVR” was further modelized with the mechanism “hindsight bias” an almost significant result (p-value ≈ 0.05) was observed and showed a lower epidemic growth of 0.06 per million of inhabitants in the country. This is a significant decrease, because it means that simply by using an environmental factor mechanism on the landing pages of their official COVID-19 websites, governments can reduce the spread of the disease and the number of cases. Inversely, when the “hindsight bias” mechanism is not used countries score high in the IVR variable, and there is a higher epidemic growth in the country.

## V. Discussion

In essentially every country on Earth, central government authorities directed and ran the response to the COVID-19 crisis which became a global pandemic within months [70]. In Europe, despite most countries being a part of the EU, all member and non-member states reacted at the national level and differently to one another. Very strict containment measures were implemented in France, Spain and Italy [71] and tighter movement controls were introduced in Germany, the Czech Republic, Slovakia, Denmark, Poland, Latvia, Lithuania, Estonia and Cyprus, sometimes going as far as completely closing their borders [72]. In the USA, the federal government lead the national response to COVID-19 and state and local health departments stood on the front lines [73]. All the decisions were taken with the same aim: to limit population movements and social life, two aggravating factors of the spread of the virus [74]. Controlled studies have shown the effectiveness of handwashing in removing viruses from the hands and as such this was promoted widely on the landing page of the governments’ official COVID-19 websites under analysis [75]. Nevertheless, countries differed in their responses as evidenced by the variation of the “look and feel” of the 33 websites assessed in this study. This includes variation in colour, in font, in the amount of text, the use of pictures and videos and so on [76]. This study sought investigate these differences further by analysing the alternative uses of behavioural insights and their impact on the number of cases and deaths of COVID-19.

The landing pages of the websites of 33 institutional governments were compared to the WHO data on the number of COVID-19 cases and deaths per million for each country. In order to address the cultural differences in each country, the six dimensions of national culture of Hofstede *et al*, were used PDI, IDV, MAS, UAI, LTO and IVR. Three levels of analysis were carried out: the individual level, the social level and the environmental level. In total, 18 mechanisms were observed, assessed and compared across the 33 countries from March 1^st^ til May 1^st^ 2020, every fifteen days.

### A behavioural consensus was observed across all 33 countries

What was observed is that there was a behavioural consensus across all 33 countries in the way they handled the sanitary crisis. As could be expected in view of the context, the majority of countries were uncomfortable with the uncertainty of the COVID-19 pandemic and displayed individualistic cultural traits rather than collective ones. Half of the countries accepted and expected that power was distributed unequally and displayed more competitiveness. Further of the 9 Individual factors that were analysed, the majority of countries implemented on average 80% of them and on average 66% of the environmental ones. Not one country used the mechanism of disgust to encourage handwashing and instead all 33 countries used social norms to do so. All 33 websites elicited emotions such as empathy and responsibility and none the feeling of fear or shame [77]. Further none of the countries used the behavioural insight of positive reinforcement or reward giving [78].

The majority (29 out of 33 countries), all provided information about the importance of handwashing and provided a hotline help number, but two countries (Lithuania and Croatia) did not provide help to support the coping of self-isolation through social networks, phone and video calls or social media or provide a hotline access. Interestingly, Norway and Switzerland adopted the exact same behaviours for all 18 mechanisms. At the individual level, two groups of respectively six and five countries used the exact same mechanisms for every action under every domain.

### Individual and Social nudges have no impact, environmental nudges can help slow the virus growth scale

There was therefore a great level of consensus in the choice of behavioural insights used. However, when looking at the impact of the use of these with the number of cases and deaths, the results show that the use of individual factors and social factors had no effect on the epidemic scale growth. Inversely, the use of environmental factors affected the growth scale. This suggests that countries applying more environmental behavioural insights demonstrated a lower a slower growth of the COVID-19 pandemic. Our results show that simply by using an environmental factor mechanism on the landing pages of their official COVID-19 websites, governments could reduce the spread of the disease and the number of cases.

These results demonstrate that irrespective of the framing of the behavioural insight depending on the culture, the use of individual and social nudges has no impact. It has been previously documented that people find it quite hard to wash their hands effectively and consistently [79] to such an extent that faecal bacteria was found on the hands of 28% of people tested in a 2008 study [80]. Lack of awareness about the effectiveness of soap, water, and scrubbing [81], practical barriers to handwashing such as difficult access to water and soap, hygiene fatigue are only a few of the reasons that have been reported to attempt to explain why people don’t wash their hands despite dozens of studies showing that it can reduce the spread of infections by around 15% to 20% [82]. Additionally, the impact of a population taking some protective measures may be negative on others and wearing a mask could make people feel sufficiently protected such that they don’t make the effort to further wash their hands [83].

### Explanation of the divergence between the observed and expected results

In this paper, the authors suggest that in the case of the COVID-19, the lack of impact of the individual and social nudges stems from the fact that there is a disconnected dissonance with reality, meaning that everybody has a different perception and experience of the threat [84]. Hand-washing can prevent life-threatening diseases, and even save lives [85], but if people are not directly exposed to the threat and don’t visually see its impact on their own family or community, they may feel an “optimism bias” that leads them to underestimate the risk [86]. An “overconfidence effect” can also be a cause for not spending the appropriate amount of time washing one’s hands thoroughly [87]. As indicated by the levels of LTO, a reluctance to change may mean people display a “status quo bias,” [88] or exhibit a “present bias,” which suggests that the short term consequences bear more importance than the long term ones [89]. Because not everyone abides by the rules, those people who are more concerned by the threat may fear being mocked, ostracised or ridiculed by their peers and this could explain why people don’t follow the guidance. The impact of religious beliefs can mean that some populations may believe that the virus is a divine punishment and therefore resign themselves and alternatively others may dismiss COVID-19 in the light of more visibly dramatic challenges such as millions of refugees living in camps [90].

Similarly, because the stay at home order were implemented reasonably quickly, the social nudges did not carry any impact because people were alone at home. Their reality of society was disconnected to what was being suggested by the governments on the landing pages of the COVID-19 websites. Inversely, the environmental nudges were impactful because people perceived themselves as an *us*, a new *we*, based on a feeling of common fate, which motivated care and concern for others [91]. Faced with danger, a new sense of social identity develops among populations that tend to focus more on the *we* rather than *me* thinking [92].

### People seek information beyond governmental websites

This study focused on the behavioural insights used on 33 governmental websites but a recent survey showed that to the question, “what is the most relied upon source of information”, only 40% answered National government sources, 34% Global health organizations like the WHO and 29%National health authorities like the CDC [93]. Deceived by governments, journalists and even scientists, people turned to social media where everything and anything was posted. People relayed false messages on how the virus spread and killed. Studies showed that the start of the lockdown was paired with a very strong activity on social networks, with high peaks and a record number of original publications, likes and retweets[94]. Undeniably social media is an extensive source of information, albeit uncurated [95]. Further a large-scale survey covering 58 countries and over 100,000 respondents between late March and early April 2020 showed that the perception of citizens was that their government and public response was weak and that fellow citizens were not doing enough [96]. People tend to overestimate small risks and underestimate large ones and as such their average confidence in their knowledge is wrongly exacerbated. This human irrationality means governmental communication tends to be paternalistic and consequently often ill-perceived by the citizens [97].

### Using Scientists as role models may encourage governmental website’s traffic

This crisis is characterized by psychological uncertainty due to the limited and potentially inconsistent answers available. Further, behavioural science shows that if the amount of information is perceived as overwhelming, people tend to disengage [98], and therefore do not receive the valuable information buried in the noise [99]. 45% of respondents to a recent survey said that “It has been difficult for me to find reliable and trustworthy information about the virus and its effects” [100]. Governments websites need to be seen as trustworthy and reliable. And since 85% of respondents of the same survey expressed the wish “to hear more from scientists and less from politicians” [101] to what extent could scientists be engaged to recommend accessing the governmental websites? As discussed previously, exposure to celebrity advocacy messages can impact issue engagement, and can have an important influence on the public’s health-related attitudes, beliefs, and behaviour [102]. Involvement of a scientist-celebrity through media exposure could effectively promote health-related messages. Scientists are coming out of their ivory tours and it is recommended to use them as role models in a society that is asking for more trustworthy public speakers.

### Designing recursive nudges will increase the impact of individual and social interventions

A nudge, in the words of economist Richard Thaler and legal scholar Cass Sunstein, is “any aspect of the choice architecture that alters people’s behaviour in a predictable way without forbidding any options or significantly changing / their economic incentives” [103]. As discussed previously, in order for a behavioural intervention to be impactful, the targeted population needs to be psychologically or physically able to undertake the behaviour and the environment needs to facilitate it [104]. Further, co-operation requires clarity, repetition and as shown in this paper a sense of collectiveness that demonstrates that the behaviour is in the group’s interest. Establishing norms and building trust encourages individuals to commit to a behaviour. A recursive nudge means that its impact will snowball over time [105]. For handwashing to become part of the collective subconscious, repetition needs to be built in over time. It starts when individuals are children…schools need to teach the importance of washing hands regularly during the day, before and after specific tasks. There are nudges that have become an integral part of our collective memory because they were both recursive and precise thereby psychologically targeting our individual or group’s behaviour [106]. The individual and social nudges that were used on the government’s websites need to be reiterated at every level of the individuals’ life, not just affixed onto a website. Permanent, visual and human nudges need to be designed [107]. This could include engaging neighbour’s in supporting each other by regular visits to enquire as to whether they are washing hands and following the governments advice. Visual cues should be omnipresent in public transports, in shops, at work and so on. The visual impact of children is also a significant psychological driver and having a little boy or little girl ring your doorbell once a week to enquire as to whether you are safe, are staying at home and washing your hands regularly would not only drive down the relevance and importance of the message but further reassert the feeling of community, which as demonstrated by these results increases abidance to the guidance [108]. It’s important that policymakers, rather than seeing groups as problems to be overcome, leverage the fact that people in groups help one another. Humans are a social species and we rely on each other for happiness and survival. It has never been more important to realize that we are stronger together [109]. Leveraging people’s intrinsic feelings of unity is a behavioural nudge that will help reduce the spread of the disease [110].

## VI. Conclusion

At the time of writing the COVID-19 crisis is still a daily concern for millions of individuals around the world and for their respective governments. Countries are starting to lift the lockdown measures, whilst others are seeing a surge in the amount of cases and deaths and scientists are discussing a putative second wave of the spread of the epidemic. This paper sought to take a snapshot at a specific time point of the behavioural nudges used by 33 governments on the landing page of their official COVID-19 websites in order to attempt to understand the behavioural response of a subset of the 7 billion individuals impacted by this pandemic. We showed that the nudges were mostly unsuccessful irrespective of their framing and suggested that in order for them to foster impact they should not only be more precise but importantly recursive. It is not enough to affix behavioural interventions on websites and expect citizens to abide by them. Further, this research has demonstrated the importance of environmental nudges and suggested that framing individual and social ones in a broader context that reemphasizes the feeling of collectiveness is a key to success. Together with the new phases of the spread of the virus will come new rules and guidance. Public health policies need to learn from the success and failures of these past few months and address behavioural change of the population on a global scale in a more targeted manner. It is hoped that this paper will provide some insight on how to do so.

## Data Availability

Data available

## VII. Acknowledgments

We declare that this manuscript is original, has not been published before and is not currently being considered for publication elsewhere.

We know of no conflicts of interest associated with this publication, and there has been no financial support for this work that could have influenced its outcome. The manuscript has been read and approved for submission by all the named authors.

## References

1. Brown GR, Dickins TE, Sear R and Laland KN, Evolutionary accounts of human behavioural diversity. Phil. Trans. R. Soc. B, 2011; 366, 313–324 doi:10.1098/rstb.2010.0267

2. Brown GR, Dickins TE, Sear R and Laland KN, Evolutionary accounts of human behavioural diversity. Phil. Trans. R. Soc. B, 2011; 366, 313–324 doi:10.1098/rstb.2010.0267

3. Brown GR, Dickins TE, Sear R and Laland KN, Evolutionary accounts of human behavioural diversity. Phil. Trans. R. Soc. B, 2011; 366, 313–313 doi:10.1098/rstb.2010.0267

4. Brown GR, Dickins TE, Sear R and Laland KN, Evolutionary accounts of human behavioural diversity. Phil. Trans. R. Soc. B, 2011; 366, 313–324 doi:10.1098/rstb.2010.0267

5. Brown GR, Dickins TE, Sear R and Laland KN, Evolutionary accounts of human behavioural diversity. Phil. Trans. R. Soc. B, 2011; 366, 313–324 doi:10.1098/rstb.2010.0267

6. Morse SS, Garwin RL, Olsiewski PJ, Next Flu Pandemic: What to Do Until the Vaccine Arrives? Science, 2006; 314:10 929

7. COVID-19 Tracker. 2020. Available from: http://www.covid19tracker.co.in/research/

8. Morse SS, Garwin RL, Olsiewski PJ, Next Flu Pandemic: What to Do Until the Vaccine Arrives? Science, 2006; 314:10929

9. Morse SS, Garwin RL, Olsiewski PJ, Next Flu Pandemic: What to Do Until the Vaccine Arrives? Science, 2006; 314:10929

10. Bavel JJV, Baicker K, Boggio PS, Capraro V, Cichocka A, Cikara M et al, Using social and behavioural science to support COVID-19 pandemic response. Nat Hum Behav, 2020; https://doi.org/10.1038/s41562-020-0884-z

11. Datta S, Responding to COVID-19 in the Developing World. Behavioral Scientist. 2020 Apr 20. Available from: https://behavioralscientist.org/responding-to-covid-19-in-the-developing-world/

12. Lunn P, Belton C, Lavin C, McGowan F, Timmons S and Robertson D, Using Behavioural Science to Help Fight the Coronavirus. Journal of Behavioral Public Administration, 2020; Vol. 3 No. 1 Available from: https://journal-bpa.org/index.php/jbpa/article/view/147

13. Barnett M, To Achieve Sustainability, We Need to Go Upstream in the Design Process. Behavioral Scientist. 2020 Apr 20. Available from: https://behavioralscientist.org/to-achieve-sustainability-we-need-to-go-upstream-in-the-design-process/

14. Barnett M, To Achieve Sustainability, We Need to Go Upstream in the Design Process. Behavioral Scientist. 2020 Apr 20. Available from: https://behavioralscientist.org/to-achieve-sustainability-we-need-to-go-upstream-in-the-design-process/

15. Barnett M, To Achieve Sustainability, We Need to Go Upstream in the Design Process. Behavioral Scientist. 2020 Apr 20. Available from: https://behavioralscientist.org/to-achieve-sustainability-we-need-to-go-upstream-in-the-design-process/

16. Lunn P, Belton C, Lavin C, McGowan, F, Timmons S and Robertson D, Using Behavioural Science to Help Fight the Coronavirus. Journal of Behavioral Public Administration, 2020; Vol. 3 No. 1 Available from: https://journal-bpa.org/index.php/jbpa/article/view/147

17. Lunn P, Belton C, Lavin C, McGowan F, Timmons S and Robertson D, Using Behavioural Science to Help Fight the Coronavirus. Journal of Behavioral Public Administration, 2020; Vol. 3 No. 1 Available from: https://journal-bpa.org/index.php/jbpa/article/view/147

18. Lunn P, Belton C, Lavin C, McGowan F, Timmons S and Robertson D, Using Behavioural Science to Help Fight the Coronavirus. Journal of Behavioral Public Administration, 2020; Vol. 3 No. 1 Available from: https://journal-bpa.org/index.php/jbpa/article/view/147

19. Lunn P, Belton C, Lavin C, McGowan F, Timmons S and Robertson D, Using Behavioural Science to Help Fight the Coronavirus. Journal of Behavioral Public Administration, 2020; Vol. 3 No. 1 Available from: https://journal-bpa.org/index.php/jbpa/article/view/147

20. Lunn P, Belton C, Lavin C, McGowan F, Timmons S and Robertson D, Using Behavioural Science to Help Fight the Coronavirus. Journal of Behavioral Public Administration, 2020; Vol. 3 No. 1 Available from: https://journal-bpa.org/index.php/jbpa/article/view/147

21. Lunn P, Belton C, Lavin C, McGowan F, Timmons S and Robertson D, Using Behavioural Science to Help Fight the Coronavirus. Journal of Behavioral Public Administration, 2020; Vol. 3 No. 1 Available from: https://journal-bpa.org/index.php/jbpa/article/view/147

22. Lunn P, Belton C, Lavin C, McGowan F, Timmons S and Robertson D, Using Behavioural Science to Help Fight the Coronavirus. Journal of Behavioral Public Administration, 2020; Vol. 3 No. 1 Available from: https://journal-bpa.org/index.php/jbpa/article/view/147

23. Lunn P, Belton C, Lavin C, McGowan F, Timmons S and Robertson D, Using Behavioural Science to Help Fight the Coronavirus. Journal of Behavioral Public Administration, 2020; Vol. 3 No. 1 Available from: https://journal-bpa.org/index.php/jbpa/article/view/147.

24. Lunn P, Belton C, Lavin C, McGowan F, Timmons S and Robertson D, Using Behavioural Science to Help Fight the Coronavirus. Journal of Behavioral Public Administration, 2020; Vol. 3 No. 1 Available from: https://journal-bpa.org/index.php/jbpa/article/view/147

25. Lunn P, Belton C, Lavin C, McGowan F, Timmons S and Robertson D, Using Behavioural Science to Help Fight the Coronavirus. Journal of Behavioral Public Administration, 2020; Vol. 3 No. 1 Available from: https://journal-bpa.org/index.php/jbpa/article/view/147

26. Bavel JJV, Baicker K, Boggio PS, Capraro V, Cichocka A, Cikara M et al, Using social and behavioural science to support COVID-19 pandemic response. Nat Hum Behav, 2020; https://doi.org/10.1038/s41562-020-0884-z

27. Bavel JJV, Baicker K, Boggio PS, Capraro V, Cichocka A, Cikara M et al, Using social and behavioural science to support COVID-19 pandemic response. Nat Hum Behav, 2020; https://doi.org/10.1038/s41562-020-0884-z

28. Lunn P, Belton C, Lavin C, McGowan F, Timmons S and Robertson D, Using Behavioural Science to Help Fight the Coronavirus. Journal of Behavioral Public Administration, 2020; Vol. 3 No. 1 Available from: https://journal-bpa.org/index.php/jbpa/article/view/147

29. Lunn P, Belton C, Lavin C, McGowan F, Timmons S and Robertson D, Using Behavioural Science to Help Fight the Coronavirus. Journal of Behavioral Public Administration, 2020; Vol. 3 No. 1 Available from: https://journal-bpa.org/index.php/jbpa/article/view/147

30. Lunn P, Belton C, Lavin C, McGowan F, Timmons S and Robertson D, Using Behavioural Science to Help Fight the Coronavirus. Journal of Behavioral Public Administration, 2020; Vol. 3 No. 1 Available from: https://journal-bpa.org/index.php/jbpa/article/view/147.

31. Lunn P, Belton C, Lavin C, McGowan F, Timmons S and Robertson D, Using Behavioural Science to Help Fight the Coronavirus. Journal of Behavioral Public Administration, 2020; Vol. 3 No. 1 Available from: https://journal-bpa.org/index.php/jbpa/article/view/147

32. Lunn P, Belton C, Lavin C, McGowan F, Timmons S and Robertson D, Using Behavioural Science to Help Fight the Coronavirus. Journal of Behavioral Public Administration, 2020; Vol. 3 No. 1 Available from: https://journal-bpa.org/index.php/jbpa/article/view/147

33. Bavel JJV, Baicker K, Boggio PS, Capraro V, Cichocka A, Cikara M et al, Using social and behavioural science to support COVID-19 pandemic response. Nat Hum Behav, 2020; https://doi.org/10.1038/s41562-020-0884-z

34. Lunn P, Belton C, Lavin C, McGowan F, Timmons S and Robertson D, Using Behavioural Science to Help Fight the Coronavirus. Journal of Behavioral Public Administration, 2020; Vol. 3 No. 1 Available from: https://journal-bpa.org/index.php/jbpa/article/view/147

35. Bavel JJV, Baicker K, Boggio PS, Capraro V, Cichocka A, Cikara M et al, Using social and behavioural science to support COVID-19 pandemic response. Nat Hum Behav, 2020; https://doi.org/10.1038/s41562-020-0884-z

36. Becker AB, Engaging celebrity? Measuring the impact of issue-advocacy messages on situational involvement, complacency and apathy. Journal Celebrity Studies, 2012, 3:2 https://doi.org/10.1080/19392397.2012.679462 Available from: https://www.tandfonline.com/doi/abs/10.1080/19392397.2012.679462

37. Becker AB, Engaging celebrity? Measuring the impact of issue-advocacy messages on situational involvement, complacency and apathy. Journal Celebrity Studies, 2012, 3:2https://doi.org/10.1080/19392397.2012.679462 Available from: https://www.tandfonline.com/doi/abs/10.1080/19392397.2012.679462

38. Bavel JJV, Baicker K, Boggio PS, Capraro V, Cichocka A, Cikara M et al, Using social and behavioural science to support COVID-19 pandemic response. Nat Hum Behav, 2020; https://doi.org/10.1038/s41562-020-0884-z

39. Bish A and Michie S, Demographic and attitudinal determinants of protective behaviours during a pandemic: A review. British Journal of Health Psychology, 2010, 15: 797–824. doi:10.1348/135910710X485826

40. Bish A and Michie S, Demographic and attitudinal determinants of protective behaviours during a pandemic: A review. British Journal of Health Psychology, 2010, 15: 797–824. doi:10.1348/135910710X485826

41. Bish A and Michie S, Demographic and attitudinal determinants of protective behaviours during a pandemic: A review. British Journal of Health Psychology, 2010, 15: 797–824. doi:10.1348/135910710X485826

42. Bavel JJV, Baicker K, Boggio PS, Capraro V, Cichocka A, Cikara M et al, Using social and behavioural science to support COVID-19 pandemic response. Nat Hum Behav, 2020; https://doi.org/10.1038/s41562-020-0884-z

43. Fetzer T, Witte M, Hensel L, Jachimowicz JM, Haushofer J, Ivchenko A et al, Global Behaviors and Perceptions at the Onset of the COVID-19 Pandemic. NBER Working Paper No. 27082 Issued in May 2020 Available from: https://www.nber.org/papers/w27082

44. Lunn P, Belton C, Lavin C, McGowan F, Timmons S and Robertson D, Using Behavioural Science to Help Fight the Coronavirus. Journal of Behavioral Public Administration, 2020; Vol. 3 No. 1 Available from: https://journal-bpa.org/index.php/jbpa/article/view/147

45. Lunn P, Belton C, Lavin C, McGowan F, Timmons S and Robertson D, Using Behavioural Science to Help Fight the Coronavirus. Journal of Behavioral Public Administration, 2020; Vol. 3 No. 1 Available from: https://journal-bpa.org/index.php/jbpa/article/view/147

46. Lunn P, Belton C, Lavin C, McGowan F, Timmons S and Robertson D, Using Behavioural Science to Help Fight the Coronavirus. Journal of Behavioral Public Administration, 2020; Vol. 3 No. 1 Available from: https://journal-bpa.org/index.php/jbpa/article/view/147

47. Lunn P, Belton C, Lavin C, McGowan F, Timmons S and Robertson D, Using Behavioural Science to Help Fight the Coronavirus. Journal of Behavioral Public Administration, 2020; Vol. 3 No. 1 Available from: https://journal-bpa.org/index.php/jbpa/article/view/147

48. Fetzer T, Witte M, Hensel L, Jachimowicz JM, Haushofer J, Ivchenko A et al, Global Behaviors and Perceptions at the Onset of the COVID-19 Pandemic. NBER Working Paper No. 27082 Issued in May 2020 Available from: https://www.nber.org/papers/w27082

49. Lunn P, Belton C, Lavin C, McGowan F, Timmons S and Robertson D, Using Behavioural Science to Help Fight the Coronavirus. Journal of Behavioral Public Administration, 2020; Vol. 3 No. 1 Available from: https://journal-bpa.org/index.php/jbpa/article/view/147

50. Lunn P, Belton C, Lavin C, McGowan F, Timmons S and Robertson D, Using Behavioural Science to Help Fight the Coronavirus. Journal of Behavioral Public Administration, 2020; Vol. 3 No. 1 Available from: https://journal-bpa.org/index.php/jbpa/article/view/147

51. Lunn P, Belton C, Lavin C, McGowan F, Timmons S and Robertson D, Using Behavioural Science to Help Fight the Coronavirus. Journal of Behavioral Public Administration, 2020; Vol. 3 No. 1 Available from: https://journal-bpa.org/index.php/jbpa/article/view/147.

52. Bish A and Michie S, Demographic and attitudinal determinants of protective behaviours during a pandemic: A review. British Journal of Health Psychology, 2010, 15: 797–824. doi:10.1348/135910710X485826

53. Bish A and Michie S, Demographic and attitudinal determinants of protective behaviours during a pandemic: A review. British Journal of Health Psychology, 2010, 15: 797–824. doi:10.1348/135910710X485826

54. Bish A and Michie S, Demographic and attitudinal determinants of protective behaviours during a pandemic: A review. British Journal of Health Psychology, 2010, 15: 797–824. doi:10.1348/135910710X485826

55. Bish A and Michie S, Demographic and attitudinal determinants of protective behaviours during a pandemic: A review. British Journal of Health Psychology, 2010, 15: 797–824. doi:10.1348/135910710X485826

56. Bish A and Michie S, Demographic and attitudinal determinants of protective behaviours during a pandemic: A review. British Journal of Health Psychology, 2010, 15: 797–824. doi:10.1348/135910710X485826

57. Beltekian D, Gavrilov D, Hasell J, Macdonald B, Mathieu E, Ortiz-Ospina E et al, Data on COVID-19 (coronavirus) by Our World in Data. owid/covid-19-data 2020. Available from: https://github.com/owid/covid-19-data/tree/master/public/data

58. Hofstede G, Hofstede GJ, Minkov M et al, National culture. Hofstede Insights. 2020. Available from: https://hi.hofstede-insights.com/national-culture

59. Cecchi-Dimeglio P, How to react to a biased performance review and prevent them in the future. HBR Guide for Women at Work, 2018, 70–84

60. Hofstede G, Hofstede GJ, Minkov M et al, National culture. Hofstede Insights. 2020. Available from: https://hi.hofstede-insights.com/national-culture

61. Hofstede G, Hofstede GJ, Minkov M et al, National culture. Hofstede Insights. 2020. Available from: https://hi.hofstede-insights.com/national-culture

62. Hofstede G, Hofstede GJ, Minkov M et al, National culture. Hofstede Insights. 2020. Available from: https://hi.hofstede-insights.com/national-culture

63. Hofstede G, Hofstede GJ, Minkov M et al, National culture. Hofstede Insights. 2020. Available from: https://hi.hofstede-insights.com/national-culture

64. Hofstede G, Hofstede GJ, Minkov M et al, National culture. Hofstede Insights. 2020. Available from: https://hi.hofstede-insights.com/national-culture

65. Hofstede G, Hofstede GJ, Minkov M et al, National culture. Hofstede Insights. 2020. Available from: https://hi.hofstede-insights.com/national-culture

66. Jia L, Li K, Jiang Y et al, Prediction and analysis of Coronavirus Disease 2019. arXiv preprint arXiv:2003.05447, 2020

67. Bates DM and Watts DG, Nonlinear Regression Analysis and Its Applications, 1988. Wiley

68. Cecchi-Dimeglio P, Designing Equality In The Legal Profession: A Nudging Approach. Harvard Negotiation Law Review, 2019; 24:1 2–26

69. Cecchi Dimeglio P and Honorable Brenneur B,(eds.) Interdisciplinary Handbook of Dispute Resolution / Manuel Interdisciplinaire sur les Modes de Prévention et Règlement des Différends: Regards Croisées. Bruylant-Larcier, 2015, pp. 1500

70. Kitson-Pantano R, To What Extent is the EU just as Federal as the USA? EuropaNova, 2020. Available from: https://www.europanova.eu/actualites/covid-19-to-what-extent-is-the-eu-just-as-federal-as-the-usa

71. Kitson-Pantano R, To What Extent is the EU just as Federal as the USA? EuropaNova, 2020. Available from: https://www.europanova.eu/actualites/covid-19-to-what-extent-is-the-eu-just-as-federal-as-the-usa

72. Kitson-Pantano R, To What Extent is the EU just as Federal as the USA? EuropaNova, 2020. Available from: https://www.europanova.eu/actualites/covid-19-to-what-extent-is-the-eu-just-as-federal-as-the-usa

73. Kitson-Pantano R, To What Extent is the EU just as Federal as the USA? EuropaNova, 2020. Available from: https://www.europanova.eu/actualites/covid-19-to-what-extent-is-the-eu-just-as-federal-as-the-usa

74. Kitson-Pantano R, To What Extent is the EU just as Federal as the USA? EuropaNova, 2020. Available from: https://www.europanova.eu/actualites/covid-19-to-what-extent-is-the-eu-just-as-federal-as-the-usa

75. Hallsworth M, Handwashing Can Stop a Virus—So Why Don’t We Do It? Behavioral Scientist. 2020 Mar 4. Available from: https://behavioralscientist.org/handwashing-can-stop-a-virus-so-why-dont-we-do-it-coronavirus-covid-19/

76. Cecchi Dimeglio P, How Gender Biais Corrupts Performance Reviews, And What To do About It. Harvard Business Review. 2017, April 2:8

77. Cecchi Dimeglio P and Honorable Brenneur B,(eds.) Interdisciplinary Handbook of Dispute Resolution / Manuel Interdisciplinaire sur les Modes de Prévention et Règlement des Différends: Regards Croisées. Bruylant-Larcier, 2015, pp. 1500

78. Cecchi Dimeglio P and Honorable Brenneur B,(eds.) Interdisciplinary Handbook of Dispute Resolution / Manuel Interdisciplinaire sur les Modes de Prévention et Règlement des Différends: Regards Croisées. Bruylant-Larcier, 2015, pp. 1500

79. Hallsworth M, Handwashing Can Stop a Virus—So Why Don’t We Do It? Behavioral Scientist. 2020 Mar 4. Available from: https://behavioralscientist.org/handwashing-can-stop-a-virus-so-why-dont-we-do-it-coronavirus-covid-19/

80. Hallsworth M, Handwashing Can Stop a Virus—So Why Don’t We Do It? Behavioral Scientist. 2020 Mar 4. Available from: https://behavioralscientist.org/handwashing-can-stop-a-virus-so-why-dont-we-do-it-coronavirus-covid-19/

81. Hallsworth M, Handwashing Can Stop a Virus—So Why Don’t We Do It? Behavioral Scientist. 2020 Mar 4. Available from: https://behavioralscientist.org/handwashing-can-stop-a-virus-so-why-dont-we-do-it-coronavirus-covid-19/

82. Hallsworth M, Handwashing Can Stop a Virus—So Why Don’t We Do It? Behavioral Scientist. 2020 Mar 4. Available from: https://behavioralscientist.org/handwashing-can-stop-a-virus-so-why-dont-we-do-it-coronavirus-covid-19/

83. Hallsworth M, Handwashing Can Stop a Virus—So Why Don’t We Do It? Behavioral Scientist. 2020 Mar 4. Available from: https://behavioralscientist.org/handwashing-can-stop-a-virus-so-why-dont-we-do-it-coronavirus-covid-19/

84. Cecchi Dimeglio P, Système de gestion de cooperation dans la période d’exploitation des Partenariats public-privé dans le secteur de la santé. Revue d’arbitrage et de médiation / Journal of Arbitration and Mediation, 2013, 35–58

85. Lopez G and Belluz J, Wash your damn hands. Vox.com. 2020 Apr 14. Available from: https://www.vox.com/2020/2/28/21157769/how-to-prevent-the-coronavirus

86. Innovation and Youth Teams at the UNDP Regional Hub for the Arab States, Nudge Lebanon, and B4 Development, Using behavioural insights to respond to COVID-19. United Nations Development Programme. 2020 May 7. Available from: https://www.undp.org/content/undp/en/home/stories/using-behavioural-insights-to-respond-to-covid-19-.html

87. Innovation and Youth Teams at the UNDP Regional Hub for the Arab States, Nudge Lebanon, and B4 Development, Using behavioural insights to respond to COVID-19. United Nations Development Programme. 2020 May 7. Available from: https://www.undp.org/content/undp/en/home/stories/using-behavioural-insights-to-respond-to-covid-19-.html respond-to-covid-19-.html

88. Innovation and Youth Teams at the UNDP Regional Hub for the Arab States, Nudge Lebanon, and B4 Development, Using behavioural insights to respond to COVID-19. United Nations Development Programme. 2020 May 7. Available from: https://www.undp.org/content/undp/en/home/stories/using-behavioural-insights-to-respond-to-covid-19-.html

89. Innovation and Youth Teams at the UNDP Regional Hub for the Arab States, Nudge Lebanon, and B4 Development, Using behavioural insights to respond to COVID-19. United Nations Development Programme. 2020 May 7. Available from: https://www.undp.org/content/undp/en/home/stories/using-behavioural-insights-to-respond-to-covid-19-.html

90. Innovation and Youth Teams at the UNDP Regional Hub for the Arab States, Nudge Lebanon, and B4 Development, Using behavioural insights to respond to COVID-19. United Nations Development Programme. 2020 May 7. Available from: https://www.undp.org/content/undp/en/home/stories/using-behavioural-insights-to-respond-to-covid-19-.html

91. Davies J, We Aren’t Selfish After All. Nautilus, 2020. Apr 29. Available from: http://nautil.us/issue/84/outbreak/we-arent-selfish-after-all?mc_cid=172dcncc2&mc_eid=0e40a038f3

92. Davies J, We Aren’t Selfish After All. Nautilus, 2020. Apr 29. Available from: http://nautil.us/issue/84/outbreak/we-arent-selfish-after-all?mc_cid=172dcncc2&mc_eid=0e40a038f3

93. Edelman Trust Barometer 2020. 2020 Available from:https://www.edelman.com/trustbarometer

94. Kitson-Pantano R, The Consequences of Poor Communication. Absolutys, 2020. Available from: https://www.absolutys.com/institute

95. Kitson-Pantano R, The Consequences of Poor Communication. Absolutys, 2020. Available from: https://www.absolutys.com/institute

96. Fetzer T, Witte M, Hensel L, Jachimowicz JM, Haushofer J, Ivchenko A et al, Global Behaviors and Perceptions at the Onset of the COVID-19 Pandemic. NBER Working Paper No. 27082 Issued in May 2020 Available from: https://www.nber.org/papers/w27082

97. Gigerenzer G, The irrationality paradox. Behavioral And Brain Sciences, 2004; 27:3336–338

98. Sagara N, Dagogo Jack T, Galak J, Behavioral Science Insights Amidst the Covid-19 Outbreak. IPSOS. 2020. Apr 6. Available from: https://www.ipsos.com/en-us/knowledge/consumer-shopper/behavioral-science-insights-amidst-the-covid19-outbreak

99. Sagara N, Dagogo Jack T, Galak J, Behavioral Science Insights Amidst the Covid-19 Outbreak. IPSOS. 2020. Apr 6. Available from: https://www.ipsos.com/en-us/knowledge/consumer-shopper/behavioral-science-insights-amidst-the-covid19-outbreak

100. Edelman Trust Barometer 2020. 2020 Available from: https://www.edelman.com/trustbarometer

101. Edelman Trust Barometer 2020. 2020 Available from: https://www.edelman.com/trustbarometer

102. Becker AB, Engaging celebrity? Measuring the impact of issue-advocacy messages on situational involvement, complacency and apathy. Journal Celebrity Studies, 2012, 3:2 https://doi.org/10.1080/19392397.2012.679462 Available from:https://www.tandfonline.com/doi/abs/10.1080/19392397.2012.679462

103. Innovation and Youth Teams at the UNDP Regional Hub for the Arab States, Nudge Lebanon, and B4 Development, Using behavioural insights to respond to COVID-19. United Nations Development Programme. 2020 May 7. Available from: https://www.undp.org/content/undp/en/home/stories/using-behavioural-insights-to-respond-to-covid-19-.html

104. Lunn P, Belton C, Lavin C, McGowan F, Timmons S and Robertson D, Using Behavioural Science to Help Fight the Coronavirus. Journal of Behavioral Public Administration, 2020; Vol. 3 No. 1 Available from: https://journal-bpa.org/index.php/jbpa/article/view/147

105. Cecchi-Dimeglio P, Diversity Dividend, MIT Press Forthcoming 2020

106. Cecchi-Dimeglio P, Designing Equality In The Legal Profession: A Nudging Approach. Harvard Negotiation Law Review, 2019; 24:12–26

107. Cecchi-Dimeglio P, Designing Equality In The Legal Profession: A Nudging Approach. Harvard Negotiation Law Review, 2019; 24:12–26

108. Cecchi Dimeglio P, Système de gestion de cooperation dans la période d’exploitation des Partenariats public-privé dans le secteur de la santé. Revue d’arbitrage et de médiation / Journal of Arbitration and Mediation, 2013, 35–58

109. Cecchi Dimeglio P, Système de gestion de cooperation dans la période d’exploitation des Partenariats public-privé dans le secteur de la santé. Revue d’arbitrage et de médiation / Journal of Arbitration and Mediation, 2013, 35–58

110. Davies J, We Aren’t Selfish After All. Nautilus, 2020. Apr 29. Available from: http://nautil.us/issue/84/outbreak/we-arent-selfish-after-all?mc_cid=172dc11cc2&mc_eid=0e40a038f3

